# Multimodal Assessment of Atrial Cardiomyopathy Identifies a High-Risk Subgroup of Embolic Stroke of Undetermined Source

**DOI:** 10.64898/2026.02.06.26345789

**Authors:** Niklas Bach, Louisa Bauermeister, Lisann Böhnke, Alexander Zuk, Stephan Salmen, Vasileios Gkizas, Alanna Ebigbo, Ralf Gold, Arash Haghikia, Michael Gotzmann

## Abstract

**Background:** Embolic stroke of undetermined source (ESUS) is associated with a high risk of recurrence. However, randomized trials have not shown superiority of anticoagulation over aspirin in unselected patients. Atrial cardiomyopathy (AtCM) may identify a high-risk ESUS phenotype. We investigated whether multimodal AtCM markers distinguish ESUS from controls and predict adverse clinical outcomes.

**Methods:** In this prospective single-center study, consecutive ESUS patients and age– and sex-matched controls without known cardiac disease were enrolled and followed for ≥12 months. All participants underwent clinical assessment, measurement of NT-proBNP, 12-lead ECG, and transthoracic echocardiography. The primary endpoint was a composite of all-cause death, recurrent stroke, transient ischemic attack, myocardial infarction, or newly detected atrial fibrillation. Cox regression analyses identified independent predictors and optimal cutoff values.

**Results:** The study included 103 ESUS patients (mean age 70.6±13.3 years) and 123 controls. Compared with controls, ESUS patients had higher NT-proBNP levels, more frequent advanced interatrial block (IAB), and impaired left atrial function. Over a mean follow-up of 470±205 days, 29 ESUS patients experienced the primary endpoint. Independent predictors were NT-proBNP >420 pg/mL, advanced IAB, E′ ≤9 cm/s, left atrial volume index ≥29 mL/m², and left atrial ejection fraction <50%. A risk score incorporating these variables identified a high-risk ESUS subgroup (≥3 factors) in which >50% experienced a cardiovascular event within 1 year.

**Conclusions:** AtCM features are common in ESUS and strongly associated with adverse outcomes. A multimodal assessment incorporating NT-proBNP, ECG, and echocardiography identifies a high-risk ESUS subgroup that merits targeted evaluation in future anticoagulation trials.

## Introduction

Ischemic stroke remains a leading cause of death and long-term disability worldwide (**1**). Approximately 15–25% of ischemic strokes are classified as embolic stroke of undetermined source (ESUS) (**2**). Despite advances in diagnostic evaluation and acute management, patients with ESUS continue to face an annual recurrence risk of ≈5%, underscoring an urgent need for more effective secondary prevention strategies (**3**).

Although ESUS is presumed to have an embolic pathophysiology, randomized trials have failed to demonstrate superiority of oral anticoagulation over aspirin for secondary prevention (**4–7**). A prespecified subgroup analysis of NAVIGATE-ESUS, however, suggested benefit from anticoagulation in patients with marked left atrial enlargement (**8**). This observation supports the hypothesis that a subset of ESUS patients may harbor an underlying atrial cardiomyopathy (AtCM) in whome anticoagulation could be therapeutically relevant.

An updated 2024 consensus statement emphasized that clinically relevant AtCM can independently predispose patients to both atrial fibrillation and ischemic stroke (**9**). Nevertheless, robust and validated diagnostic thresholds for AtCM remain lacking, and it is unknown whether patients with AtCM who are in sinus rhythm benefit from anticoagulation. A recent clinical consensus statement from the Heart Failure Association of the European Society of Cardiology conceptualizes AtCM as a progressive disease spectrum ranging from healthy atria to atrial failure, characterized by structural, electrical, and functional remodeling that may promote thromboembolism even in the absence of atrial fibrillation (**10**).

The ARCADIA trial recently evaluated anticoagulation in ESUS patients selected for AtCM markers, including elevated N-terminal pro–B-type natriuretic peptide (NT-proBNP), left atrial enlargement, and abnormal terminal P-wave force in lead V1 (PTFV1). However, anticoagulation did not reduce recurrent stroke compared with aspirin in this population (**7**). Our prior work suggests that the ARCADIA selection criteria and cut-off values may have lacked sufficient specificity to accurately capture clinically relevant AtCM (**11**). Thus, a key question remains unresolved: does a distinct ESUS subgroup with AtCM exist that carries a persistently increased risk of recurrence and may benefit from anticoagulation? Given the substantial residual risk after ESUS, this issue is of major clinical importance (**12,13**).

Multiple diagnostic modalities may reflect the presence of AtCM (**14**), but electrocardiography and echocardiography are particularly scalable for population-level phenotyping.

Accordingly, this study had two objectives. First, we compared AtCM-related biomarkers and phenotypes (NT-proBNP, P-wave indices, and left atrial hemodynamics) between ESUS patients and age– and sex-matched controls without overt cardiac disease. Second, we sought to identify predictors defining a high-risk ESUS subgroup with elevated recurrence risk. Improved phenotyping of such patients may inform the design of future anticoagulation trials in ESUS.

## Methods

### Study Design and Population

This prospective, single-center study represents an analysis of the ARCANA study (AtRial CArdiomyopathy – prevaleNce And clinical impact), conducted through interdisciplinary collaboration between cardiology/electrophysiology, neurology, and internal medicine at St. Josef Hospital Bochum, Ruhr University Bochum.

From July 2022 to December 2024, consecutive patients with ESUS were enrolled. An age-and sex-matched control group without prior stroke or transient ischemic attack (TIA) and without manifest cardiac disease served as comparators. All participants were followed for at least 12 months. Written informed consent was obtained from all patients. The study was approved by the local ethics committee (registration number 22-7535).

### Inclusion and Exclusion Criteria

**ESUS Group:** Inclusion criteria were: 1) ESUS according to current criteria (**15**): imaging-confirmed nonlacunar ischemic infarction, no ≥50% stenosis in an artery supplying the infarct territory, no major cardioembolic source, and no alternative specific stroke etiology; 2) exclusion of a patent foramen ovale; 3) sinus rhythm at study inclusion; 4) age ≥18 and <90 years; 5) life expectancy >1 year; 6) written informed consent.

Exclusion criteria were: 1) atrial fibrillation before or during hospitalization; 2) structural heart disease (e.g., severe valvular disease or LVEF <30%); 3) severe renal failure (GFR <15 mL/min); 4) severe hepatic disease; 5) manifest hyperthyroidism; 6) prior valve replacement or coronary artery bypass surgery; 7) continuous pacemaker stimulation.

**Control Group:** Controls were age– and sex-matched individuals with: 1) no known or manifest heart disease (except left ventricular hypertrophy due to hypertension); 2) no history of stroke or TIA.

**General Exclusion Criteria (All Patients):** 1) Severe valvular disease; 2) use of antiarrhythmic drugs; 3) prior pulmonary vein isolation or cardiac surgery; 4) continuous pacemaker stimulation or non-sinus rhythm; 5) electrical or pharmacological cardioversion within 3 months; 6) end-stage renal disease; 7) LVEF <30%.

### Clinical and Laboratory Assessment

Medical history was obtained in all patients, and blood samples were collected and analyzed. Blood pressure was measured immediately before echocardiography. Twelve-lead ECGs were recorded immediately before or after echocardiography.

### ECG and Echocardiography

The methodology has been described previously in the ARCANA study (**11**). Briefly, ECGs were recorded at 50 mm/s and 10 mm/mV. Two blinded investigators independently analyzed all ECGs; discrepancies were resolved by a third reviewer. P-wave parameters were assessed according to International Society of Electrocardiology recommendations (**16**). Heart rate, P-wave duration, PR interval, and QRS duration were recorded. Advanced interatrial block was defined as P-wave duration ≥120 ms with biphasic morphology in leads II, III, and aVF (Figure 1).

**Figure 1.**
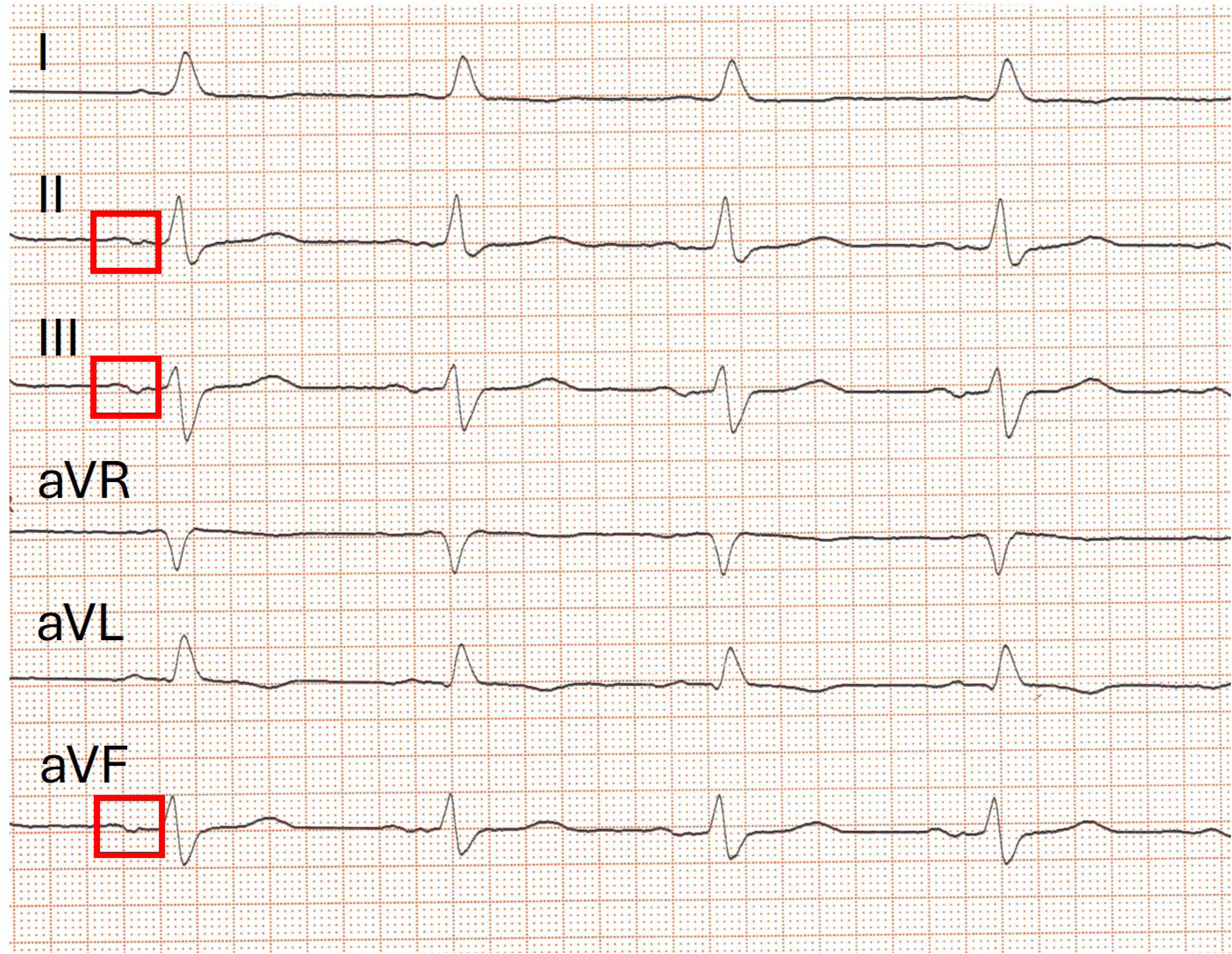
Advanced interatrial block on 12-lead ECG. Representative 12-lead electrocardiogram demonstrating advanced interatrial block. The P wave is highlighted in red boxes, illustrating prolonged P-wave duration with biphasic morphology in the inferior leads, consistent with advanced interatrial block.

Transthoracic echocardiography was performed by an experienced investigator according to European Society of Cardiology guidelines (**17,18**) using Vivid E9 or E95 systems (GE Healthcare, Horten, Norway). Images were archived and analyzed with EchoPAC software (GE Healthcare). The PA-TDI interval was measured from the onset of the P wave to the peak late diastolic velocity at the lateral mitral annulus (**19**). Tissue Doppler parameters (E′, A′, S′) were obtained in the apical four-chamber view at lateral and septal sites and averaged. Left atrial strain was assessed in apical two– and four-chamber views (**20**).

### Management of ESUS Patients

All ESUS patients were treated in a comprehensive stroke unit. Diagnosis was made by experienced neurologists based on current recommendations. Treatment and secondary prevention were independent of the study protocol and included aspirin 100 mg daily. No patient received oral anticoagulation.

### Outcomes

Events were defined as: 1) all-cause death; 2) recurrent stroke; 3) new TIA; 4) myocardial infarction; 5) newly detected atrial fibrillation. The primary endpoint was the composite of these five events. The secondary endpoint was the composite of all-cause death and recurrent stroke.

### Follow-Up

Patients were contacted annually by telephone. For reported events (stroke, TIA, myocardial infarction, atrial fibrillation), the date of diagnosis was recorded and verified by medical documentation whenever possible. For deceased patients, information was obtained from relatives and primary care physicians.

### Statistical Analysis

Statistical analyses were performed using SPSS version 26. Continuous variables are presented as mean ± SD. Between-group comparisons were conducted using the unpaired t test for normally distributed variables and the Mann–Whitney U test or Kruskal–Wallis test for non-normally distributed variables, as appropriate. Categorical variables were compared using the χ² test.

Time to first event was used for endpoint analyses. First, outcomes were compared between ESUS patients and controls. The cumulative incidence of the primary and secondary endpoints was estimated using the Kaplan–Meier method, and survival curves were compared with the log-rank test.

In a second analysis, ESUS patients with and without the primary endpoint were compared. All variables were first examined in univariable analyses. Parameters significantly associated with the primary endpoint were entered into Cox proportional hazards regression models to identify independent predictors. Receiver operating characteristic (ROC) analyses were performed to determine optimal cutoff values. Identified risk factors were then combined into a composite risk model, categorizing patients according to the presence of <3 versus ≥3 factors. Event-free survival across risk strata was assessed using Kaplan–Meier curves and compared with the log-rank test. A two-sided P<0.05 was considered statistically significant.

## Results

### Study Population

Between July 2022 and December 2024, 1,622 patients with ischemic stroke were treated at our hospital. During this period, all consecutive patients fulfilling ESUS criteria were screened for eligibility. A total of 112 patients met the prespecified inclusion and exclusion criteria. Four patients declined participation and five were lost to 1-year follow-up. The final ESUS cohort comprised 103 patients (50 women, 53 men; mean age 70.6 ± 13.3 years). The control group included 123 individuals (59 women, 64 men; mean age 70.0 ± 14.6 years). ESUS and control groups did not differ with respect to age, sex, body mass index, or left ventricular ejection fraction (Table 1). Mean follow-up was 470 ± 205 days.

**Table 1.**
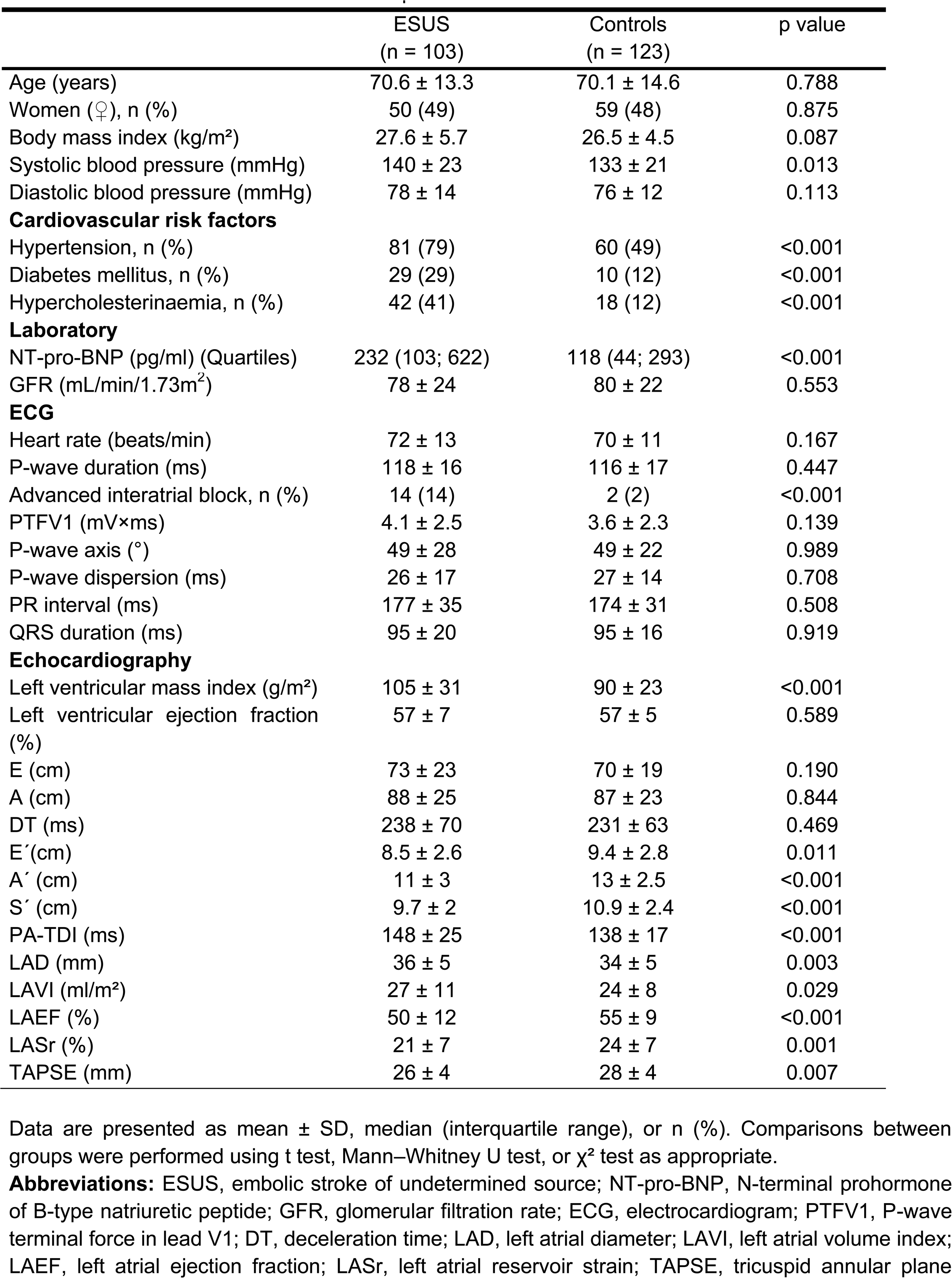
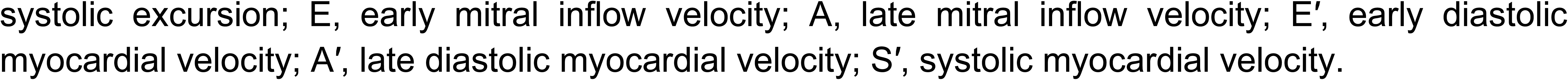
Baseline characteristics of ESUS patients and matched controls.

Compared with controls, ESUS patients had a higher prevalence of cardiovascular risk factors, higher NT-proBNP levels, a greater frequency of advanced interatrial block, and significant differences in multiple echocardiographic parameters, particularly those reflecting diastolic function and left atrial hemodynamics (Table 1).

### Clinical Events

During follow-up, patients in the control group experienced few events: death (n=2), stroke (n=0), transient ischemic attack (TIA; n=1), myocardial infarction (n=0), and newly detected atrial fibrillation (n=1). In contrast, events occurred significantly more frequently among ESUS patients (P<0.001), including death (n=6), recurrent stroke (n=11), TIA (n=8), myocardial infarction (n=2), and newly detected atrial fibrillation (n=7). Four patients experienced two events (stroke and TIA, n=2; stroke and atrial fibrillation, n=1; stroke and death, n=1); for the primary endpoint, only the first event was considered. Event rates at 1 year and over the entire study period are presented in Table 2.

**Table 2.** Clinical events during the first year and over the entire study follow-up.

|  | ESUS<br>(n =<br>103) | Controls<br>(n = 123) |
| --- | --- | --- |
| All-cause death | 5/6 | 2/2 |
| Stroke* | 9/11 | 0 |
| Transitory ischemic attack | 7/8 | 1/1 |
| Myocardial infarction | 2/2 | 0 |
| Atrial fibrillation | 5/7 | 1/1 |

Data are shown as number of events during the first year / total events during follow-up. Four ESUS patients experienced two events (stroke and TIA, n = 2; stroke and atrial fibrillation, n = 1; stroke and death, n = 1); for the primary endpoint, the first event was analyzed.
Abbreviations: ESUS, embolic stroke of undetermined source; TIA, transient ischemic attack.

Notably, 11 ESUS patients sustained recurrent ischemic stroke during follow-up. In 9 cases, the recurrent event again fulfilled ESUS criteria, whereas 1 was classified as small-vessel disease, 1 as large-artery atherosclerosis, and 1 as stroke of undetermined etiology. In 7 ESUS patients, clinical atrial fibrillation was newly diagnosed during follow-up, with a mean time to diagnosis of 328±221 days after study enrolment.

Kaplan–Meier curves illustrate the cumulative incidence of the composite primary endpoint (death, stroke, TIA, myocardial infarction, and atrial fibrillation; Figure 2A) and the secondary endpoint (death and stroke; Figure 2B) in both groups.

**Figure 2.**
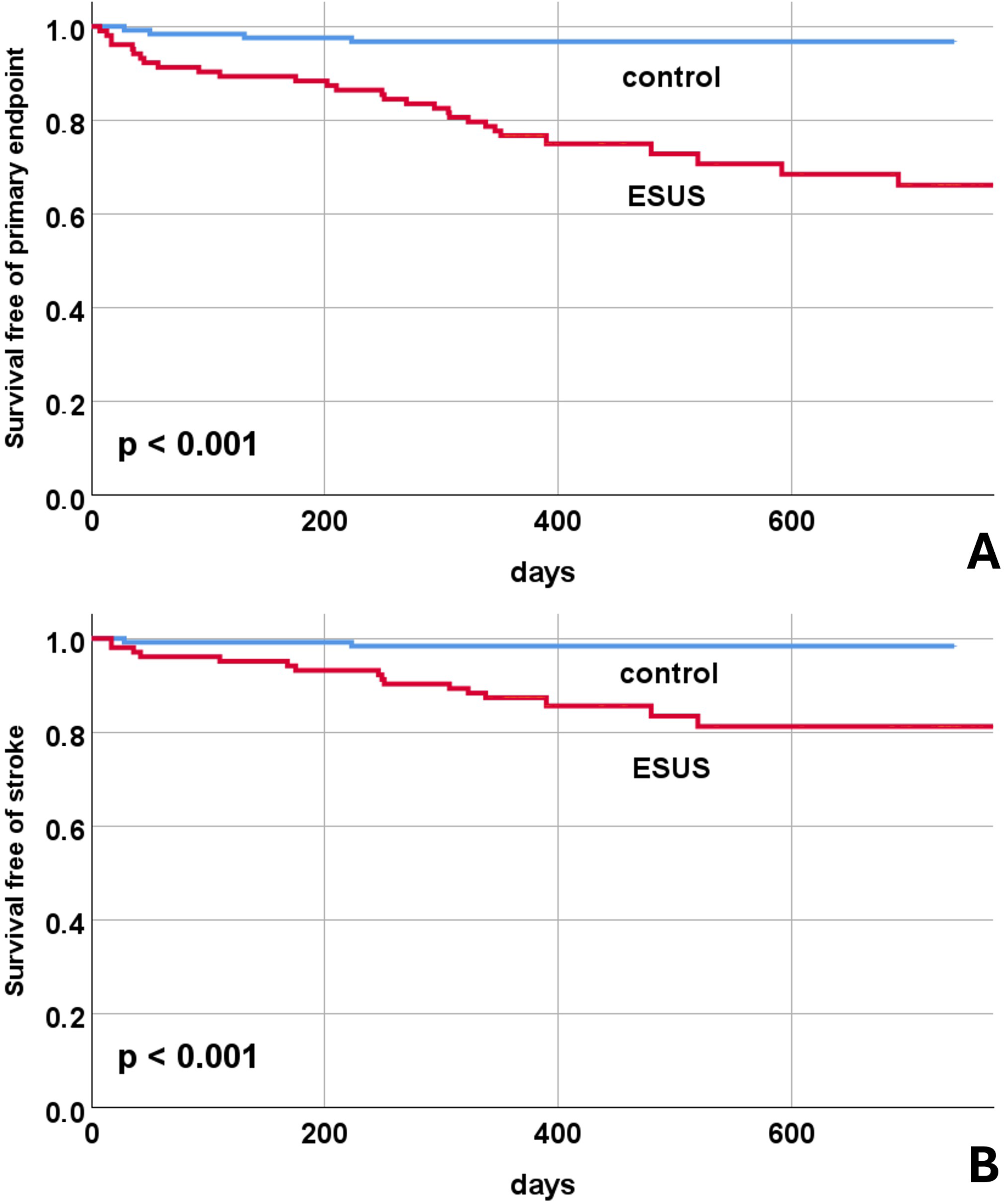
Kaplan–Meier curves for ESUS patients and matched controls. (A) Cumulative event-free survival for the primary endpoint, defined as the composite of all-cause death, recurrent stroke, transient ischemic attack (TIA), myocardial infarction, and newly detected atrial fibrillation. (B) Cumulative event-free survival for the secondary endpoint, defined as all-cause death and recurrent stroke. The ESUS group had significantly higher event rates compared with controls (log-rank test, p < 0.001 for both endpoints). **Abbreviations:** ESUS, embolic stroke of undetermined source; TIA, transient ischemic attack.

### Predictors of the Primary Endpoint in ESUS Patients

Overall, 29 ESUS patients reached the primary endpoint. Compared with those without events, patients with events were older, had a higher burden of cardiovascular risk factors, higher NT-proBNP levels, and significant differences in several hemodynamic and atrial functional parameters (Table 3).

**Table 3:** ESUS patients with and without primary study endpoint.

|  | Primary study<br>endpoint<br>(n = 29) | No primary study<br>endpoint<br>(n = 74) | p value |
| --- | --- | --- | --- |
| Age (years) | 76 ± 14 | 68 ± 12 | 0.008 |
| Women (♀), n (%) | 18 (62) | 32 (43) | 0.097 |
| Body mass index (kg/m<sup>2</sup>) | 27.3 ± 6.4 | 27.7 ± 5.5 | 0.751 |
| Systolic blood pressure (mmHg) | 146 ± 23 | 138 ± 22 | 0.095 |
| Diastolic blood pressure (mmHg) | 80 ± 15 | 78 ± 14 | 0.482 |
| **Cardiovascular risk factors** |  |  |  |
| Hypertension, n (%) | 27 (93) | 54 (74) | 0.031 |
| Diabetes mellitus, n (%) | 12 (41) | 17 (23) | 0.062 |
| Hypercholesterinaemia, n (%) | 15 (52) | 27 (36) | 0.172 |
| **Laboratory** |  |  |  |
| NT-pro-BNP (pg/ml) (Quartiles) | 650 (197; 2291) | 176 (91; 346) | <0.001 |
| GFR (mL/min/1.73m<sup>2</sup>) | 73 ± 28 | 81 ± 22 | 0.159 |
| **ECG** |  |  |  |
| Heart rate (beats/min) | 72 ± 13 | 72 ± 13 | 0.952 |
| P-wave duration (ms) | 119 ± 18 | 117 ± 15 | 0.533 |
| Advanced interatrial block, n (%) | 11 (38) | 3 (4) | <0.001 |
| PTFV1 (mV×ms) | 3.8 ± 3.2 | 4.1 ± 2.2 | 0.509 |
| P-wave axis (°) | 51 ± 37 | 49 ± 25 | 0.705 |
| P-wave dispersion (ms) | 25 ± 17 | 26 ± 17 | 0.764 |
| PR interval (ms) | 187 ± 46 | 173 ± 29 | 0.068 |
| QRS duration (ms) | 95 ± 19 | 95 ± 20 | 0.939 |
| **Echocardiography** |  |  |  |
| Left ventricular mass index (g/m<sup>2</sup>) | 112 ± 35 | 103 ± 29 | 0.215 |
| Left ventricular ejection fraction (%) | 57 ± 8 | 57 ± 7 | 0.902 |
| E (cm) | 81 ± 29 | 70 ± 19 | 0.029 |
| A (cm) | 91 ± 30 | 86 ± 23 | 0.411 |
| DT (ms) | 245 ± 75 | 235 ± 68 | 0.497 |
| E' (cm) | 7.6 ± 1.7 | 8.9 ± 2.8 | 0.004 |
| A' (cm) | 10 ± 2.8 | 12 ± 2.9 | 0.017 |
| S' (cm) | 9.2 ± 2.1 | 9.9 ± 2 | 0.099 |
| PA-TDI (ms) | 160 ± 29 | 143 ± 22 | 0.002 |
| LAD (mm) | 37 ± 5 | 35 ± 5 | 0.112 |
| LAVI (ml/m<sup>2</sup>) | 33 ± 12 | 24 ± 9 | <0.001 |
| LAEF (%) | 46 ± 15 | 52 ± 10 | 0.017 |
| LASr (%) | 18 ± 7 | 22 ± 7 | 0.014 |
| TAPSE (mm) | 26 ± 4 | 26 ± 4 | 0.629 |

Data are presented as mean ± SD, median (interquartile range), or n (%). Comparisons between groups were performed using t test, Mann–Whitney U test, or $\chi^2$ test as appropriate.
**Abbreviations:** ESUS, embolic stroke of undetermined source; NT-pro-BNP, N-terminal prohormone of B-type natriuretic peptide; GFR, glomerular filtration rate; ECG, electrocardiogram; PTFV1, P-wave terminal force in lead V1; DT, deceleration time; LAD, left atrial diameter; LAVI, left atrial volume index; LAEF, left atrial ejection fraction; LASr, left atrial reservoir strain; TAPSE, tricuspid annular plane systolic excursion; E, early mitral inflow velocity; A, late mitral inflow velocity; E', early diastolic myocardial velocity; A', late diastolic myocardial velocity; S', systolic myocardial velocity.

Variables significantly associated with the primary endpoint in univariable analyses were entered into Cox proportional hazards regression models to identify independent predictors. NT-proBNP, advanced interatrial block, E′, left atrial volume index (LAVI), and left atrial ejection fraction (LAEF) emerged as independent predictors of the primary endpoint.

Receiver operating characteristic (ROC) analyses identified optimal cutoff values for NT-proBNP and echocardiographic parameters. For NT-proBNP, the area under the curve (AUC) was 0.746 (95% CI, 0.627–0.866; P<0.001), with an optimal cutoff >420 pg/mL (sensitivity 62%, specificity 78%). For E′, the AUC was 0.649 (95% CI, 0.541–0.757; P=0.007), with a cutoff ≤9 cm/s (sensitivity 76%, specificity 50%). For LAVI, the AUC was 0.735 (95% CI, 0.627–0.843; P<0.001), with a cutoff ≥29 mL/m² (sensitivity 64%, specificity 77%). For LAEF, the AUC was 0.655 (95% CI, 0.518–0.792; P=0.027), with a cutoff <50% (sensitivity 67%, specificity 61%). Corresponding hazard ratios are presented in Table 4.

**Table 4.**
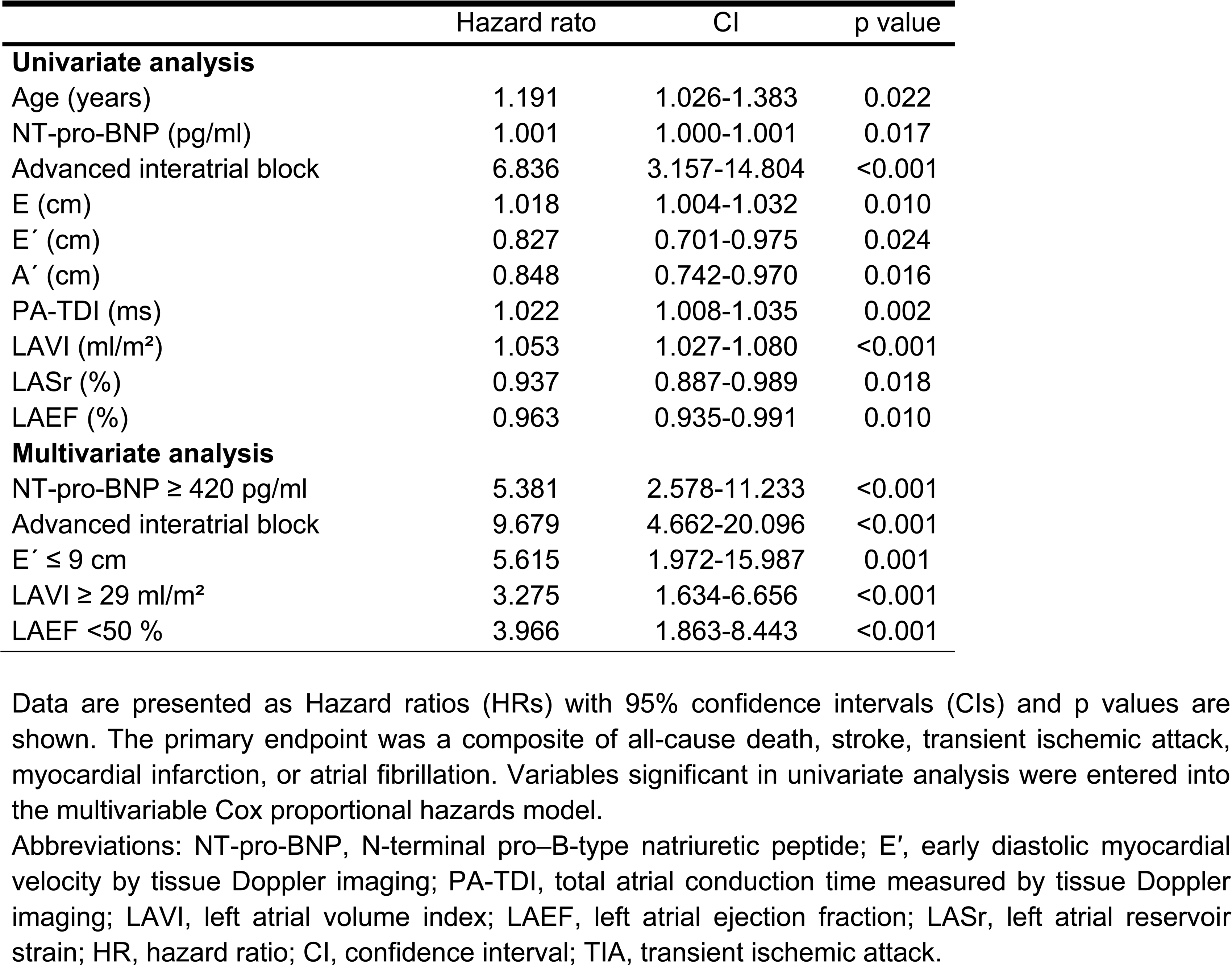
Univariate and multivariate Cox regression analyses for predictors of the primary study endpoint.

### Risk Stratification

Based on the five independent predictors, a risk score was derived classifying patients into groups with <3 (n = 75), or ≥3 risk factors (n = 28). Kaplan–Meier curves demonstrate increasing event rates across risk strata for the primary endpoint (Figure 3A) and the secondary endpoint (Figure 3B). In the high-risk group (≥3 risk factors; 27% of all patients), 17 of 29 primary endpoint events and 10 of 16 secondary endpoint events occurred.

**Figure 3.**
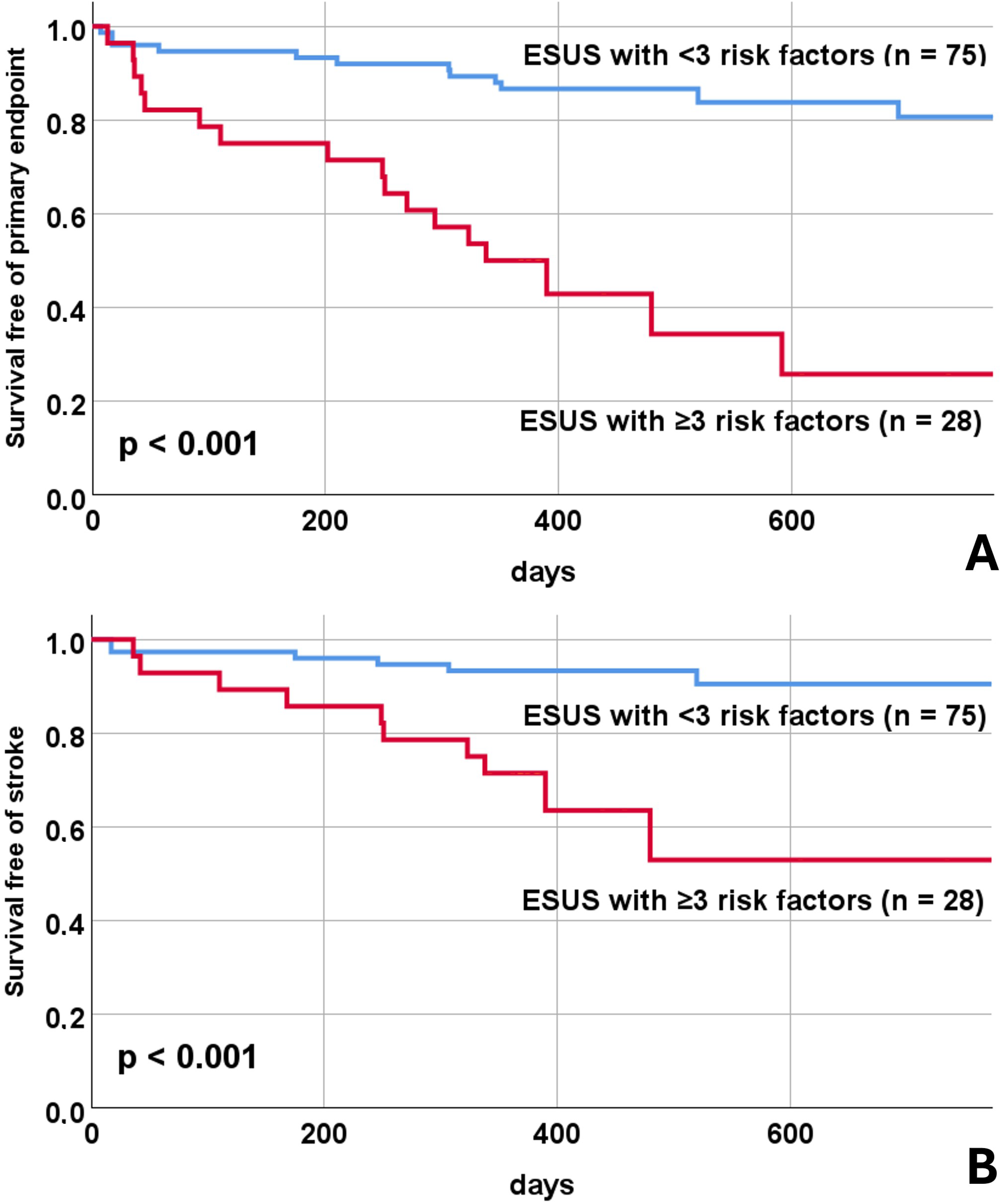
Kaplan–Meier curves stratified by risk score in ESUS patients. (A) Event-free survival for the primary endpoint (composite of all-cause death, recurrent stroke, transient ischemic attack, myocardial infarction, and newly detected atrial fibrillation) stratified by the number of independent risk factors: <3 vs ≥3. (B) Event-free survival for the secondary endpoint (all-cause death and recurrent stroke) stratified by risk score. Patients with ≥3 risk factors (high-risk group) had significant higher event rates compared to patients with <3 risk factors (log-rank test, p < 0.001 for both endpoints). **Abbreviations:** ESUS, embolic stroke of undetermined source.

## Discussion

This prospective study yields several key findings. First, patients with ESUS in our cohort had a substantial risk of recurrent stroke (9% within the first year) and a high 1-year mortality (5%). Second, compared with age– and sex-matched controls, ESUS patients differed significantly in multiple parameters of left atrial anatomy and hemodynamics, supporting the presence of an underlying AtCM in a relevant subset. Third, independent predictors of clinical events were elevated NT-proBNP, advanced interatrial block (IAB), and impaired left atrial haemodynamic function. Fourth, the combination of ≥3 risk factors identified a high-risk ESUS subgroup in which >50% experienced a major cardiovascular event (death, stroke, TIA, myocardial infarction, or atrial fibrillation) within one year (Figure 3).

### ESUS and Atrial Cardiomyopathy

The entity of ESUS likely comprises a heterogeneous group of conditions requiring differentiated secondary prevention strategies (**2**). This heterogeneity may partly explain why prior randomized trials failed to demonstrate a benefit of oral anticoagulation in unselected ESUS populations.

AtCM may represent a particularly relevant pathogenic mechanism. A subgroup of the NAVIGATE-ESUS trial suggested benefit from anticoagulation among patients with marked left atrial enlargement (**8**). However, only 5% of patients met this criterion with a left atrial diameter >4.6 cm, limiting the clinical utility for broader ESUS populations.

The AtCM concept has recently been refined with recommendations for diagnosis and grading (**9**). Prior studies indicate that AtCM is associated with ischemic stroke, possibly independent of atrial fibrillation (**14,21**). Nevertheless, validated thresholds that reliably identify clinically relevant AtCM and justify altered therapy remain lacking.

The ATTICUS trial enrolled ESUS patients with risk markers including CHA₂DS₂-VASc ≥4, atrial high-rate episodes, left atrial enlargement >45 mm, spontaneous echo contrast or low left atrial appendage flow velocity, or patent foramen ovale. Although some of these criteria overlap with AtCM markers, apixaban did not reduce stroke recurrence compared with aspirin (**6**). Similarly, the ARCADIA trial selected patients using AtCM-related criteria (PTFV1 ≥5000 μV·ms, NT-proBNP ≥250 pg/mL, or indexed left atrial diameter ≥3 cm/m²), yet anticoagulation was not superior to aspirin (**7**). Notably, most patients were enrolled based solely on NT-proBNP ≥250 pg/mL, a nonspecific marker. Only 1% qualified based on indexed left atrial size, and we recently demonstrated that PTFV1 may not reliably identify AtCM (**11**). Thus, whether ARCADIA truly studied an AtCM-enriched population remains uncertain.

### Clinical Implications of Atrial Cardiomyopathy in ARCANA

The recent HFA–ESC consensus statement by Weerts et al. emphasizes AtCM as a continuum from subclinical atrial remodeling to overt atrial failure, integrating structural, electrical, and functional domains as key drivers of thromboembolic risk independent of atrial fibrillation (**10**).

Our findings extend this framework by demonstrating that ESUS patients with a multimodal AtCM phenotype—defined by elevated NT-proBNP, advanced interatrial block, and impaired echocardiographic atrial function—represent a clinically distinct subgroup with a markedly increased risk of recurrent events.

In our study, patients with ESUS exhibited significant alterations in left atrial structure and function compared with age– and sex-matched controls, including prolonged PA-TDI intervals, increased left atrial size and volume, reduced left atrial ejection fraction and reservoir strain, impaired myocardial velocities (E′, A′, S′), and increased left ventricular mass index. In addition, ESUS patients more frequently demonstrated advanced IAB and elevated NT-proBNP levels. Collectively, these findings support a relevant cardiac—and specifically atrial—substrate contributing to stroke in a substantial subset of ESUS patients.

Importantly, ESUS patients who experienced clinical events during follow-up had higher NT-proBNP levelvs, more frequent advanced IAB, and more pronounced abnormalities in diastolic function and left atrial mechanics (Table 3). These observations underscore the prognostic relevance of AtCM features in ESUS and suggest that atrial dysfunction is closely linked to recurrent vascular and arrhythmic events.

In multivariable analysis, NT-proBNP levels, advanced IAB, E′ velocity, left atrial volume index (LAVI), and left atrial ejection fraction (LAEF) were independently associated with the primary endpoint. Although NT-proBNP is not specific for AtCM (**14**), it is a robust marker of myocardial stress and remodeling and served as a key selection criterion in ARCADIA. The higher cut-off identified in our cohort may improve specificity for clinically relevant AtCM and more accurately delineate a truly high-risk ESUS subgroup.

Advanced IAB reflects abnormal left atrial conduction and is closely linked to mechanical dysfunction. It is a recognized risk marker for atrial fibrillation, ischemic stroke, and left atrial thrombus formation (**10, 22–24**). We have previously demonstrated that advanced IAB is the ECG parameter most strongly associated with AtCM (**11**), and in the present study it emerged as the strongest independent predictor of adverse clinical events.

Among echocardiographic parameters, indices of left atrial structure and function differed consistently between patients with and without events. LAVI has been associated with both incident atrial fibrillation and recurrent stroke in ESUS populations, supporting its role as a marker of AtCM and risk stratification (**25**). Likewise, reduced LAEF, reflecting impaired atrial contractile function, has been linked to higher rates of recurrent ischemic stroke and new-onset atrial fibrillation in stroke cohorts without diagnosed AF (**26**), highlighting its potential value as a functional AtCM marker.

Finally, E′ ≤9 cm/s emerged as an independent predictor of outcome, reflecting impaired left ventricular relaxation and elevated left atrial filling pressures—mechanisms that may promote atrial remodeling and thrombogenicity (**27**). Taken together, these findings indicate that a multimodal AtCM phenotype encompassing biomarkers, ECG, and echocardiographic measures identifies ESUS patients at particularly high risk for recurrent events. Thus, ARCANA provides prospective clinical evidence that application of the AtCM concept in ESUS can identify patients with “atrial failure–like” characteristics who may benefit from targeted secondary prevention strategies.

### Risk Stratification

Using the five independent predictors, we derived a risk score. Notably, ≈25% of ESUS patients had ≥3 risk factors, and >50% of them had a major event within 12 months. Because NT-proBNP, advanced IAB, and altered left atrial hemodynamics reflect AtCM-related pathology, these findings strongly support the clinical relevance of AtCM in ESUS.

Our data suggest that a multimodal approach incorporating biomarkers (NT-proBNP), ECG (advanced IAB), and echocardiography (E′, LAVI, LAEF) can identify a high-risk ESUS subgroup in whom anticoagulation might merit reconsideration despite prior neutral trials.

### Limitations

This study has several limitations. First, it is a single-center investigation with a moderate sample size, which may limit generalizability. However, the cohort was prospectively assembled using strict inclusion and exclusion criteria to minimize competing stroke mechanisms. Second, echocardiographic and ECG measurements were performed at a single time point; therefore, dynamic changes in atrial structure and function over time could not be assessed. Finally, although the proposed risk score showed strong discrimination in this cohort, external validation in independent populations is required before clinical implementation.

### Conclusions

AtCM appears to underlie stroke in a relevant subset of ESUS patients and is associated with adverse clinical outcomes. A multimodal phenotype—NT-proBNP >420 pg/mL, advanced IAB, E′ velocity ≤9 cm/s, LAVI ≥ 29 ml/m² and LAEF <50 %—identifies a high-risk ESUS subgroup with markedly increased 1-year event rates. These findings may inform future trials evaluating anticoagulation in carefully selected ESUS patients with evidence of AtCM.

## Data Availability

The data that support the findings of this study are available from the corresponding author upon reasonable request. Due to the sensitive nature of the clinical data and to protect patient privacy, the data are not publicly available.

## Disclosures / Conflicts of Interest

Niklas Bach, Louisa Bauermeister, Lisann Böhnk and Alexander Zuk, Alanna Ebigbo, Stephan Salmen and Ralf Gold report that there are no conflicts of interest relevant to this manuscript.

Dr. Vasileios Gkizas reports consulting fees, honoraria, and/or speaker fees from Boehringer Ingelheim, Pfizer and Biotronik

Dr. Arash Haghikia reports consulting fees, and/or speaker fees from Novo Nordisk, Boehringer Ingelheim, Bristol Myers Squibb, Bayer, Amgen, Novartis, and Pfizer. He reports no other conflicts of interest relevant to this manuscript.

Dr. Michael Gotzmann reports consulting fees, honoraria, and/or speaker fees from Alnylam, Boehringer Ingelheim, Bristol Myers Squibb, Eli Lilly, Novartis, and Pfizer. He reports no other conflicts of interest relevant to this manuscript.

## Funding

No specific funding was received for this study.

## Abbreviations List

AtCM: atrial cardiomyopathy
ESUS: embolic stroke of undetermined source
ECG: electrocardiogram
E′: early diastolic myocardial velocity
IAB: interatrial block
LA: left atrial
LAVI: left atrial volume index
LAEF: left atrial ejection fraction
LVEF: left ventricular ejection fraction
NT-proBNP: N-terminal pro–B-type natriuretic peptide
PA-TDI: P-wave to A′ tissue Doppler interval
PTFV1: terminal P-wave force in lead V1
ROC: receiver operating characteristic
TIA: transient ischemic attack

## Notes

### Clinical Trial

This study is a prospective observational cohort study without any intervention. Therefore, registration in an ICMJE-approved clinical trials registry was not required.

### Funding Statement

This work was not supported by any external funding. The authors and their institutions did not receive any payment or services from a third party for any aspect of the submitted work, including study design, data collection, analysis, interpretation, or manuscript preparation.

### Author Declarations

The Ethics Committee of the Ruhr-University Bochum approved the study protocol (approval number: 22-7535) and all patients provided written informed consent.

## References

1 Prabhakaran S, Gonzalez NR, Zachrison KS, Adeoye O, Alexandrov AW, Ansari SA, et al; Peer Review Committee. 2026 guideline for the early management of patients with acute ischemic stroke: a guideline from the American Heart Association/American Stroke Association. Stroke. 2026;57:xxx–xxx. doi:10.1161/STR.0000000000000513

2 Ntaios G, Baumgartner H, Doehner W, Donal E, Edvardsen T, Healey JS, et al. Embolic strokes of undetermined source: a clinical consensus statement of the ESC Council on Stroke, the European Association of Cardiovascular Imaging and the European Heart Rhythm Association of the ESC. Eur Heart J. 2024;45:1701–1715.

3 Sposato LA, Sur NB, Katan M, Johansen MC, De Marchis GM, Caso V, et al. Embolic stroke of undetermined source: new data and new controversies on cardiac monitoring and anticoagulation. Neurology. 2024;103:e209535.

4 Hart RG, Sharma M, Mundl H, Kasner SE, Bangdiwala SI, Berkowitz SD, et al; NAVIGATE ESUS Investigators. Rivaroxaban for stroke prevention after embolic stroke of undetermined source. N Engl J Med. 2018;378:2191–2201.

5 Diener HC, Sacco RL, Easton JD, Granger CB, Bernstein RA, Uchiyama S, et al; RE-SPECT ESUS Steering Committee and Investigators. Dabigatran for prevention of stroke after embolic stroke of undetermined source. N Engl J Med. 2019;380:1906–1917.

6 Geisler T, Keller T, Martus P, Poli K, Serna-Higuita LM, Schreieck J, et al; ATTICUS Investigators. Apixaban versus aspirin for embolic stroke of undetermined source. NEJM Evid. 2024;3:EVIDoa2300235.

7 Kamel H, Longstreth WT Jr, Tirschwell DL, Kronmal RA, Marshall RS, Broderick JP, et al; ARCADIA Investigators. Apixaban to prevent recurrence after cryptogenic stroke in patients with atrial cardiopathy: the ARCADIA randomized clinical trial. JAMA. 2024;331:573–581.

8 Healey JS, Gladstone DJ, Swaminathan B, Eckstein J, Mundl H, Epstein AE, et al. Recurrent stroke with rivaroxaban compared with aspirin according to predictors of atrial fibrillation: secondary analysis of the NAVIGATE ESUS randomized clinical trial. JAMA Neurol. 2019;76:764–773.

9 Goette A, Corradi D, Dobrev D, Aguinaga L, Cabrera JA, Chugh SS, et al. Atrial cardiomyopathy revisited: evolution of a concept. Europace. 2024;26:euae204.

10 Weerts J, Tica O, Aranyo J, Basile C, Borizanova-Petkova A, Borovac JA, et al. Atrial cardiomyopathy: from healthy atria to atrial failure. Eur J Heart Fail. 2025;27:2173–2194. doi:10.1002/ejhf.3782

11 Kazantzi M, Mohr A, Schneider R, Labedi A, Bach N, Rößler J, et al. Association of P-wave parameters with left atrial hemodynamics in atrial cardiomyopathy. Ann Noninvasive Electrocardiol. 2026;31:e70145. doi:10.1111/anec.70145

12 Tandon K, Tirschwell D, Longstreth WT Jr, Smith B, Akoum N. Embolic stroke of undetermined source correlates to atrial fibrosis without atrial fibrillation. Neurology. 2019;93:e381–e387.

13 Kühnlein P, Mahnkopf C, Majersik JJ, Wilson BD, Mitlacher M, Tirschwell D, et al. Atrial fibrosis in embolic stroke of undetermined source: a multicenter study. Eur J Neurol. 2021;28:3634–3639.

14 Kreimer F, Gotzmann M. Left atrial cardiomyopathy: a challenging diagnosis. Front Cardiovasc Med. 2022;9:942385.

15 Diener HC, Easton JD, Hart RG, Kasner S, Kamel H, Ntaios G. Review and update of the concept of embolic stroke of undetermined source. Nat Rev Neurol. 2022;18:455–465.

16 Chen LY, Ribeiro ALP, Platonov PG, Cygankiewicz I, Soliman EZ, Gorenek B, et al. P wave parameters and indices: a critical appraisal. Circ Arrhythm Electrophysiol. 2022;15:e010435.

17 Lang RM, Badano LP, Mor-Avi V, Afilalo J, Armstrong A, Ernande L, et al. Recommendations for cardiac chamber quantification by echocardiography in adults. Eur Heart J Cardiovasc Imaging. 2015;16:233–270.

18 Badano LP, Kolias TJ, Muraru D, Abraham TP, Aurigemma G, Edvardsen T, et al. Standardization of left atrial, right ventricular, and right atrial deformation imaging. Eur Heart J Cardiovasc Imaging. 2018;19:591–600.

19 Müller P, Weijs B, Bemelmans NMAA, Mügge A, Eckardt L, Crijns HJGM, et al. Echocardiography-derived total atrial conduction time. Clin Res Cardiol. 2021;110:1734–1742.

20 Voigt JU, Mălăescu GG, Haugaa K, Badano L. How to do LA strain. Eur Heart J Cardiovasc Imaging. 2020;21:715–717.

21 Sajeev JK, Kalman JM, Dewey H, Cooke JC, Teh AW. The atrium and embolic stroke. JACC Clin Electrophysiol. 2020;6:251–261.

22 Kreimer F, Backhaus JF, Krogias C, Pflaumbaum A, Mügge A, Gotzmann M. P-wave parameters and thrombi in the left atrial appendage. Clin Cardiol. 2023;46:397–406.

23 Martínez-Sellés M, Elosua R, Ibarrola M, de Andrés M, Díez-Villanueva P, Bayés-Genis A, et al. Advanced interatrial block and stroke. Europace. 2020;22:1001–1008.

24 O’Neal WT, Kamel H, Zhang ZM, Chen LY, Alonso A, Soliman EZ. Advanced interatrial block and ischemic stroke. Neurology. 2016;87:352–356.

25 Jordan K, Yaghi S, Poppas A, Chang AD, Mac Grory B, Cutting S, et al. Left atrial volume index and ESUS. Stroke. 2019;50:1997–2001.

26 Friderichsen LBH, Larsen BS, Aplin M, Høst N, Hadad R, Christensen LM, et al. Left atrial cardiomyopathy and stroke recurrence. Int J Cardiovasc Imaging. 2026;xx:xxx–xxx. doi:10.1007/s10554-026-03629-5

27 Yaramada P, Pai RG. Markers of atrial myopathy. Echocardiography. 2016;33:177–178.

